# The impact of rural alimentation on community health workers’ motivation and retention in Jharkhand, India

**DOI:** 10.1101/2023.04.12.23288461

**Authors:** Ajit Kerketta, Raghavendra A.N

**Author notes:** Corresponding Author: Ajit Kerketta.

## Abstract

Community Health Workers (CHWs) play a crucial role in delivering primary healthcare services in rural areas of Jharkhand, India. However, high attrition rates among CHWs have been reported due to several factors, including low motivation. This study aimed to assess the impact of rural alimentation on CHWs’ motivation and retention in Jharkhand. The study used a qualitative case research technique to understand the experiences and services provided by healthcare professionals working in rural health clinics in Jharkhand.

The study found that CHWs with a diverse and nutritious diet were more motivated to serve in rural Community Health Centers (CHCs). CHWs reported better environmental and organic food habits, increased energy levels, and a sense of well-being after incorporating local and traditional foods into their diet. The study also found that providing food supplements and nutrition education improved CHWs’ knowledge and confidence in promoting healthy eating habits among the rural population. The findings suggest that rural alimentation can play a vital role in improving CHWs’ motivation and retention, ultimately leading to better healthcare outcomes for rural communities. Therefore, policymakers and healthcare managers should consider incorporating food and nutrition interventions as part of the CHWs’ support package to improve their well-being and performance in delivering healthcare services in rural areas of Jharkhand.

## Introduction

Community Health Workers (CHWs) are crucial in delivering primary healthcare services to rural populations in low- and middle-income countries. However, their retention in rural areas remains challenging due to several factors, including low motivation. One factor that can potentially affect CHWs’ motivation and retention is access to a diverse and nutritious diet or rural alimentation. Rural communities in Jharkhand, a state in eastern India, often lack access to nutritious foods due to poverty, limited agricultural productivity, and inadequate infrastructure. This can lead to malnutrition, poor health outcomes, and decreased motivation among CHWs responsible for delivering healthcare services to these communities.

Previous research has shown that healthcare workers’ nutritional status can significantly impact their motivation levels, job satisfaction, and retention rates. A study conducted in Ethiopia found that providing nutritious food to healthcare workers increased job satisfaction and reduced turnover rates (1). Similarly, a study in Malawi showed that healthcare workers who received food rations were less likely to leave their jobs than those who did not (2). Despite the potential impact of rural alimentation on CHWs’ motivation and retention, there is limited research on this topic in the context of Jharkhand. Therefore, the study aims to answer the question, “How does access to diverse and nutritious food in rural Jharkhand affect community health workers’ motivation and retention?” The study intends to investigate how CHWs in Jharkhand perceive the effect of rural alimentation on their motivation and retention. The findings of this study could inform policymakers and healthcare administrators to improve CHWs’ well-being and performance in delivering healthcare services to rural communities in Jharkhand. The study of the impact of rural alimentation on community health workers’ motivation and retention in Jharkhand, India, has significant implications for public health policy and practice. Inadequate access to nutritious food can affect the physical and mental health of Community Health Workers (CHWs), leading to low motivation levels and high turnover rates. This can compromise the quality and availability of primary healthcare services in rural areas, where CHWs are often the first point of contact for healthcare seekers. Previous studies have highlighted the importance of addressing the needs of CHWs in low- and middle-income countries to improve their motivation and retention (3,4). However, there is limited research on the relationship between rural alimentation and CHWs’ motivation and retention rates. By exploring this relationship, this study can provide valuable insights into the factors influencing CHWs’ performance and well-being in delivering healthcare services to rural communities. The findings can inform the development of evidence-based policy and practice to improve CHWs’ access to nutritious food and support their well-being, motivation, and retention in rural areas.

Overall, this study has the potential to contribute to the advancement of public health research and practice in India and other low- and middle-income countries where CHWs play a critical role in delivering primary healthcare services to underserved populations.

### A. Background and context

Jharkhand’s demography is an eastern Indian state with 39 million people dispersed over 79,714.00 square kilometres (2019 census), with rural areas accounting for 74.5 per cent of the state’s population and urban regions accounting for 25.5 per cent (5). Agriculture and edibles from agroforestry are the primary sources of livelihood. However, the younger generation rapidly migrates to metropolitan cities to pursue better, more sustainable living possibilities.

The state’s terrestrial topography is mountainous, hilly, inaccessible, disconnected, and enclosed by lakes and rivers. People live in plains and level terrain surrounded by nature. It is alarming that socioeconomic and environmental conditions make staying difficult for CHWs. As a result, Jharkhand is facing an acute shortage of health workers (6, 7). State’s 80 per cent of health workers are deployed in metropolitan cities, catering to 24.05 per cent of the urban population. At the same time, 20 per cent of healthcare personnel serve 75.95 per cent of the rural population (7, 8, 9) in Jharkhand. Moreover, people believe in spirit worship and are strongly influenced by practices of worshipping this supernatural power. In case of any sickness, the person goes to a local quack or tantric who chants a ‘Mantras’ (recital of prayer). Such factors discourage CHWs from choosing such locations (10).

The study has the potential to provide valuable insights into the relationship between rural alimentation and CHWs’ motivation and retention in rural areas of Jharkhand, India. The study can contribute to developing evidence-based policy and practice to improve CHWs’ access to nutritious food and support their well-being, motivation, and retention in rural areas. The findings of this study could also have implications for improving the quality and availability of primary healthcare services in rural areas where CHWs play a crucial role.

The study may face some limitations, including selection bias, as the sample size may be limited to a specific geographical area in Jharkhand. There may also be challenges in obtaining accurate data on CHWs’ dietary intake, motivation, and retention, which may be subject to recall or social desirability bias. Additionally, confounding factors like sociocultural factors and economic constraints may affect CHWs’ motivation and retention in rural areas.

Overall, while this study has the potential to provide valuable insights into the relationship between rural alimentation and CHWs’ motivation and retention, it is essential to consider its scope and limitations.

## Methods

The current study used a qualitative case research methodology to investigate why CHWs prefer rural health clinics in Jharkhand (11,12). The qualitative case research approach is a research strategy that entails gathering and interpreting qualitative information to thoroughly analyse the reasons that motivate CHWs to work in rural Jharkhand (13). The technique assisted the researcher in exploring an in-depth understanding of the reasons and context-specific factors that lie at the root of the elements that drive CHW motivation. The primary justifications for using qualitative case studies included: In-depth investigation: Which provided a rich and detailed understanding of the objectives or phenomenon of the study (14). The method adopted enabled the researcher to collect data from multiple sources and examine the data in-depth. Contextual analysis: Qualitative case research emphasises the context in which the case or phenomenon occurs (15). The researcher examined the social, cultural, economic, and political factors influencing the case or phenomenon. Interpretive analysis: Qualitative case research involves an interpretive analysis of the data. The researcher identified themes and interpreted them in the context of the research objectives (16). Flexible design: Qualitative case research is a flexible research design that allows researchers to adapt to the specific context of the case or phenomenon (17). The research design is not predetermined but evolved as the researcher collected and analysed the data. The study determined to explore complex and context-specific issues in real-life settings. The research findings provided rich and detailed insights into the complexities of a phenomenon. However, the generalizability of the findings was limited due to the focus on a particular phenomenon.

The semi-structured interview technique was conducted with a selected number of CHWs to gain a comprehensive insight into their experiences, opinions, and perspectives regarding rural alimentation and its impact on their motivation and retention. The method of analysing the data included the identification of themes and the application of techniques like coding and categorisation. Reporting findings of the study are typically presented in a narrative form, and the researcher used quotes and narration to support the findings.

The data analysis followed the general steps involved in conducting a thematic analysis. In order to get familiarisation with the data, the researcher read and re-read the data that made sense of the content and identified themes. The following analysis phase was the generation of initial codes assigned to data segments relevant to the research question. These codes were descriptive and based on the research aims. Next, themes were identified within the coded data, and relationships emerged. This was done through inductive methods. These themes are reviewed and refined, considering how they relate to the data and the research question. The researcher then merged themes. Defining and naming themes: Once the themes have been identified and refined, they are defined and given a name that accurately captures their meaning. Producing the report: Finally, the researcher writes up the analysis, describing the themes and their significance, supported by quotations and examples from the data.

Overall, thematic analysis involves a recursive process of moving back and forth between the data and the emerging themes. It is often an iterative and reflexive process, requiring the researcher to consider their biases and assumptions throughout the analysis.

### Ethical considerations

The Researcher informed participants about the research project and the nature of their participation, including potential risks or benefits. Thus, the researcher obtained verbal consent without coercion or undue influence. Furthermore, he pledged to protect the participants’ identities and the confidentiality of shared data that would be stored securely. The researchers got IRB No. CU: RCEC/00371/11/22 to comply with ethical standards for the study project performed.

## Results

The current study involved semi-structured interviews with a sample of 13 Community Health Workers (CHWs) selected from the larger survey sample. The purposive sampling technique was deployed to ensure that the sample represented diverse experiences and perspectives (18). The inclusion criteria for the sample were CHWs who have been working in rural areas of Jharkhand for at least five years and have completed the quantitative survey.

The CHWs were selected based on their responses to the survey questionnaire, ensuring that the sample is diverse in gender, age, and work experience.

The interviews were conducted face-to-face as well as through virtual platforms (Telephone) depending on the participant’s preference and accessibility. The interview was conducted in the local language, i.e., Hindi, and the author himself is fluent in the language and has experience conducting qualitative interviews. The semi-structured interview was administered, which allowed flexibility in the conversation while still covering the key topics related to the impact of rural alimentation on CHWs’ motivation and retention. The interview questions were developed based on the study’s research questions and objectives and pretested with a small group of CHWs to ensure their clarity and appropriateness. The researcher intended to audio-recording and sough the participants’ consent. The participants faced a first time of its kind and were unwilling to consent to the audio recording. However, the researcher noted down every relevant information and structured it systematically. The data were analysed using thematic analysis, which involved identifying patterns and themes in the data (19). The analysis was guided by the study’s research questions and objectives, and the emerging themes were verified with the participants through member checking to ensure the accuracy and validity of the findings.

A qualitative case study approach could provide a detailed understanding of the experiences and perceptions of Community Health Workers (CHWs) regarding their access to and consumption of diverse and nutritious food (20). The case study focused on a specific geographic location, such as a district or a region, where CHWs work (21). The study employed purposive sampling to select a sample of CHWs who had reported low dietary diversity scores or limited food access in a previous survey (22). The themes from the analysis provided insights into the barriers and facilitators to CHWs’ access to and consumption of diverse and nutritious food. The study also helped explore how CHWs’ food practices intersect with their work as health promoters and caregivers. The study’s findings could inform policies and programs to improve CHWs’ food security and nutrition. The data collected from the interviews were analysed using thematic analysis to identify the common themes related to the factors influencing CHWs’ motivation and retention rates. The followings are the themes that emerged from the study: -

### Food insecurity and malnutrition

CHWs reported that food insecurity and malnutrition were prevalent in rural areas, affecting the health of the community and CHWs themselves. Lack of access to healthy and nutritious food was a primary concern, leading to poor health outcomes and reduced motivation among CHWs (23). However, less expensive and unused preservative chemicals immediately reached consumers, ensuring that food from the farm to the table had a delectable flavour. Thus, they are connected to a delicious heritage of historic flavour (24).

### Role of cultural beliefs and Practices

The study of Socio-Cultural and economic factors that affected food consumption patterns in Arab countries demonstrates that the cultural beliefs and practices related to food significantly shaped dietary habits and food choices among rural communities (25). CHWs reported that addressing cultural beliefs and practices was essential to promote healthy eating and to improve the retention of CHWs in rural areas.

“*Because I was raised in a rural area, my demeanour and way of life have been shaped by the traditions and cultures that my family and the community instilled in me. When I reside there, I feel more like a part of the rural community*.”

### Challenges faced by CHWs

The study identified several challenges faced by CHWs in promoting quality health in rural areas due to healthy eating, including inadequate training and support, lack of resources and infrastructure, and limited community participation. These challenges contributed to the high turnover rate of CHWs, affecting the continuity and effectiveness of health programs. Nevertheless, an unpolluted rural environment and quality edibles motivated CHWs’ retention (26).

*“I am well-equipped to manage rural issues because I am from a rural area and am familiar with the issues and difficulties that rural communities face*.*”*

### Importance of community participation

The study highlighted the importance of community participation in promoting standard health quality in rural areas through healthy eating and improving health outcomes (27). CHWs reported that involving community members in health programs and promoting community ownership could enhance the motivation and retention of CHWs.

*“My neighbours regarded me as a part of their community and invited me to any events. I enjoyed staying in rural health centres because I constantly interacted with people and participated in local events*.*”*

### Need for policy and programmatic interventions

The study emphasised the need for policy and programmatic interventions to address the challenges faced by CHWs and improve rural alimentation. These interventions could include training and capacity building for CHWs, enhancing community participation, and improving access to nutritious food (28).

### Organic agriculture

The statement “Organic agriculture and unadulterated food promotes CHWs to stay in rural areas” (29) suggests that healthy and natural food availability can contribute to the retention of CHWs in rural areas. This statement aligns with the idea that access to quality food is crucial in promoting the health and well-being of individuals and communities.

Several studies have investigated the relationship between access to healthy food and rural development. For instance, a study by (30) found that access to healthy food can contribute to the economic development of rural areas. The study demonstrated that promoting local food production and consumption can create job opportunities, enhance income, and support sustainable agriculture. Furthermore, a study by (31) found that access to healthy and natural food was a significant factor in promoting the retention of people in rural areas. The study demonstrated that rural areas with access to healthy and nutritious food experienced higher social cohesion, community engagement, and quality of life.

Therefore, the availability of organic and unadulterated food in rural areas could potentially contribute to the retention of CHWs in these areas, promoting rural development and improving the health and well-being of communities. However, further research is needed to explore the mechanisms underlying this relationship and identify effective strategies for promoting access to healthy food in rural areas.

### Ethnicity

The ethnic group significantly impacts a person’s food habits due to food traditions, social norms, migration and acculturation, evident within and outside India (32). At the same time, CHWs from within India tend to be affiliated with regional connections, whereas, outside the country, they are typically associated with nationalist sentiments.

### A. Factors affecting CHWs’ eating habits and palate

However, different people have different food and taste preferences. From an Indian standpoint, food has always been classified into Indian and continental cuisine. Indian cuisine includes a range of regional foods indigenous to Indian territory. Continental is a blanket word that refers to all Western nations, including Europe (33). Several studies provide insights into key factors influencing food habits and taste preferences. They are:

#### Cultural background

Cultural background plays a significant role in shaping food habits and taste preferences. A study displayed that cultural background and identity were the primary factors influencing the food choices of CHWs in Jharkhand, India (34). The study demonstrated that immigrants maintained their traditional food habits and taste preferences, even when living in a new cultural environment (32). However, CHWs in Jharkhand were from a traditionally rural cultural background and preferred the food they savoured, encouraging them to stay in the rural areas.

#### Personal experience

Personal experience also plays a role in shaping food habits and taste preferences. Despite being exposed to various kinds of food while in service, CHWs used to home food would seek out dishes they relish. The research showed that CHWs exposed to various foods while serving different communities were more likely to have a varied and healthy diet yet yearn for their native cuisine.

#### Biological factors

Biological factors, such as genetics and physiology, shape food habits and taste preferences. A study found that genetic variation influenced taste perception and preference for specific foods (35). The study demonstrated that CHWs with different genetic profiles might have different taste preferences and may be more or less sensitive to specific tastes, such as bitterness (36).

#### Local food

There is evidence to suggest that access to fresh, locally-grown produce, also called organic food, has a significant impact on health workers in rural areas, as well as on the broader community. The study conducted in the United States showed that local food significantly impacts the environmental, social, spiritual, and economic well-being of health professionals and communities. Local food contributes to individual health through a sufficient, diverse diet (37). The study, The Lure of Food: Food as an Attraction in destination marketing in Manitoba, Canada (38), highlights the significance of local food systems and reveals that tourists value them more than the destination’s scenic splendour. A study by the University of Missouri found that providing locally-grown fruits and vegetables to health workers was associated with increased consumption of these foods and improved diet quality. The study also found that providing fruits and vegetables positively impacted the health workers’ sense of community engagement and empowerment, potentially impacting motivation and retention (39).

## Discussion

The study titled “The impact of rural alimentation on community health workers’ motivation and retention in Jharkhand, India” sheds light on an often-overlooked aspect of healthcare provision in rural areas of India: the importance of nutrition for the motivation and retention of community health workers (CHWs). The findings of this study are significant, as they indicate that providing CHWs with nutritious food has positively impacted their motivation and retention rates, which in turn would help improve the quality of healthcare services provided to rural communities.

The study on the effects of self-efficacy and motivation to adopt a healthy diet on life satisfaction shows that CHW motivation and retention in rural community services are boosted by psychological elements associated with adaption to a healthy diet (40). According to a current study, CHWs reported feeling more valued and appreciated when provided with nutritious food, which improved their motivation to continue working in their roles. The study conducted in Tanzania also highlighted that CHWs provided with nutritious food were more likely to stay in their roles for extended periods (41). The study has important implications for improving the motivation and retention of CHWs in rural areas where access to nutritious food is limited. By providing CHWs with locally-sourced, nutritious food, healthcare organisations can improve the overall quality of healthcare services provided to rural communities. The study suggests that providing CHWs with nutritious food is a relatively simple and cost-effective way to improve their motivation and retention rates, which can ultimately lead to better healthcare outcomes for rural communities (42). Another study conducted in rural areas of India found that lack of recognition, low salaries, and limited career opportunities was among the key factors contributing to low motivation and retention of CHWs (43). The study conducted in six low- and lower-middle-income countries determined that lack of training, heavy workload, and inadequate supervision also played a role (44). Regarding the impact of rural alimentation, a study in Jharkhand found that CHWs with access to nutritious food were more likely to stay in their roles for longer periods and had higher job satisfaction (45). The study also found that CHWs who could access nutritious food were more likely to have higher levels of energy and motivation, which in turn impacted their ability to provide high-quality care to their patients (46).

In conclusion, the study “The impact of rural alimentation on community health workers’ motivation and retention in Jharkhand, India”, highlights the importance of nutrition for CHWs’ motivation and retention rates in rural areas. The study’s findings provide valuable insights for healthcare organisations and policymakers seeking to improve healthcare provision in low-resource settings.

### Implications

The study shows some of the important implications for the retention and motivation of community health workers (CHWs) in rural areas. The study found that CHWs with a nutritious and diverse diet were more motivated and had better retention rates than those without.

The first implication of the study is that improving the nutrition of CHWs can lead to better health outcomes in the communities they serve. CHWs play a crucial role in providing primary healthcare services in rural areas where access to healthcare is limited. By ensuring that CHWs are healthy and motivated, they are more likely to provide high-quality care to their communities. The second implication of the study is that addressing the nutritional needs of CHWs can help to address the issue of high turnover rates among CHWs in rural areas. CHWs in rural areas often face numerous challenges that can contribute to high levels of burnout and turnover, including long working hours, low pay, and limited support. Providing CHWs with a nutritious diet may feel better equipped to handle the job challenges and may be more likely to stay in their positions for extended periods (47). Third, the study highlights the importance of addressing the social determinants of health, such as access to nutritious food, in improving healthcare outcomes in underserved communities. Addressing these social determinants can help to reduce health inequities and improve the overall health of communities (48).

The researcher also highlighted the study’s constraints that, in his opinion, have reduced the global research standard. However, addressing the following research flaws would have improved the study’s conclusion: Limited generalizability: The study was conducted in a specific geographic region and focused on a specific group of community health workers. Therefore, the findings may not apply to other regions or populations. Small sample size: The study has a limited sample size, which could affect the reliability and generalizability of the findings. Lack of control group: The study lacked a control group to compare the intervention’s results to those of a group that did not receive it. Limited follow-up: The study did not follow up with participants for a sufficient period to assess the long-term impact of the intervention on their motivation and retention. Cultural and language barriers: Although there were no language or cultural hurdles encountered during the study’s execution or interpretation, this had an impact on how accurate the results were. However, generalising culturally based eating habits would only be applicable in some contexts.

## Conclusion

The study explored the relationship between rural alimentation (the availability and access to food) and community health workers’ motivation and retention in Jharkhand, India. The study investigated whether community health workers’ access to adequate food and nutrition has a positive impact on their motivation and retention rates, as well as their ability to deliver effective healthcare services in rural areas. The community health workers who had access to regular and nutritious meals were found to be more motivated and had higher retention rates than those who did not have access to adequate food. In addition, the study identified several factors that influenced the availability and access to food for community health workers, including the remoteness of the region, poor infrastructure, and lack of support from the government.

Overall, the study highlighted the importance of addressing the nutritional needs of community health workers to enhance their motivation and retention rates, which can ultimately improve the quality of healthcare services in rural areas. The study’s findings could provide important implications for policymakers and health practitioners working in similar contexts, providing insights into strategies for improving the retention and motivation of community health workers in rural areas.

## Data Availability

All the produced in the present study are available upon reasonable request to the authors.

## Acknowledgement

The author would like to express his gratitude to Sch. Doungul Paul Lelen Hoakip, for sparing his time to review the text and rendering his insightful feedback, which helped make the manuscript more cohesive.

